# Simple Scoring Tool to Estimate Risk of Hospitalization and Mortality in Ambulatory and Emergency Department Patients with COVID-19

**DOI:** 10.1101/2021.02.22.21252171

**Authors:** Brandon J. Webb, Nicholas M. Levin, Nancy Grisel, Samuel M. Brown, Ithan D. Peltan, Emily S. Spivak, Mark Shah, Eddie Stenehjem, Joseph Bledsoe

**Author notes:** Corresponding Author / Reprints: Brandon J. Webb, MD, Division of Epidemiology and Infectious Diseases, Intermountain Medical Center, 5121 S. Cottonwood Drive, Murray, UT 84157.

## Abstract

**Background:** Accurate methods of identifying patients with COVID-19 who are at high risk of poor outcomes has become especially important with the advent of limited-availability therapies such as monoclonal antibodies. Here we describe development and validation of a simple but accurate scoring tool to classify risk of hospitalization and mortality.

**Methods:** All consecutive patients testing positive for SARS-CoV-2 from March 25-October 1, 2020 within the Intermountain Healthcare system were included. The cohort was randomly divided into 70% derivation and 30% validation cohorts. A multivariable logistic regression model was fitted for 14-day hospitalization. The optimal model was then adapted to a simple, probabilistic score and applied to the validation cohort and evaluated for prediction of hospitalization and 28-day mortality.

**Results:** 22,816 patients were included; mean age was 40 years, 50.1% were female and 44% identified as non-white race or Hispanic/Latinx ethnicity. 6.2% required hospitalization and 0.4% died. Criteria in the simple model included: age (0.5 points per decade); high-risk comorbidities (2 points each): diabetes mellitus, severe immunocompromised status and obesity (body mass index≥30); non-white race/Hispanic or Latinx ethnicity (2 points), and 1 point each for: male sex, dyspnea, hypertension, coronary artery disease, cardiac arrythmia, congestive heart failure, chronic kidney disease, chronic pulmonary disease, chronic liver disease, cerebrovascular disease, and chronic neurologic disease. In the derivation cohort (n=16,030) area under the receiver-operator characteristic curve (AUROC) was 0.82 (95% CI 0.81-0.84) for hospitalization and 0.91 (0.83-0.94) for 28-day mortality; in the validation cohort (n=6,786) AUROC for hospitalization was 0.8 (CI 0.78-0.82) and for mortality 0.8 (CI 0.69-0.9).

**Conclusion:** A prediction score based on widely available patient attributes accurately risk stratifies patients with COVID-19 at the time of testing. Applications include patient selection for therapies targeted at preventing disease progression in non-hospitalized patients, including monoclonal antibodies. External validation in independent healthcare environments is needed.

## Introduction

COVID-19 is a systemic infection caused by a novel betacoronavirus, SARS-CoV-2.(1) A relatively conserved set of clinical and demographic factors are now recognized to correlate with an increased risk for severe disease requiring hospitalization, mechanical ventilation and death.(2–4) Accurate methods of risk stratifying ambulatory patients at the point of test positivity have many possible applications, including prioritizing patients at highest risk of hospitalization for early treatments aimed to prevent progression to severe disease, such as monoclonal antibodies, which are both limited in availability and also more likely to be effective in high-risk groups. Several risk classification models have been proposed.(3,5–9) We describe development and validation of a simple scoring model to predict hospitalization and mortality in a large cohort of ED and ambulatory patients with COVID-19.

## Methods

Intermountain Healthcare is an integrated healthcare system that provides care to more than 1.5 million patients each year in Utah and bordering communities. As part of a systemwide COVID-19 response, Intermountain provides SARS-CoV-2 testing at 32 urgent care facilities, 23 emergency departments, and 16 community drive-up testing sites. During the study period, only polymerase chain reaction (PCR) assays were performed (Thermofisher, Waltham, MA; Cepheid, Sunnyvale, CA; Quidel, San Diego, CA, BioFire, Salt Lake City, UT; Roche, Basel, Switzerland). All testing required an order entered in the electronic health record (EHR) (Cerner, Kansas City, KS) by the ordering clinician through a structured form that requires the clinician to input the patient’s clinical symptoms and epidemiological features. These data are stored in the Intermountain Prospective Observational COVID-19 (IPOC) database, and the enterprise data warehouse. This analysis was approved by the Institutional Review Board at Intermountain Healthcare under #1051342.

We queried the IPOC database for consecutive adult patients with positive SARS-CoV-2 tests from March 25-October 1, 2020. Symptom data were extracted from the electronic test order form while demographic and co-morbidity data were obtained from the IPOC database and data warehouse using the Charlson and Elixhauser definitions (10,11). We defined immunosuppression as: recipient of a solid organ or hematopoietic stem cell transplant, on chemotherapy, biologic or other immunosuppressive agents targeting B or T cell activity, chronic corticosteroids at a prednisone-equivalent dose of 20mg per day or greater for more than 30 days, human immunodeficiency virus complicated by acquired immunodeficiency syndrome (AIDS), heritable immunodeficiency. We defined obesity as body mass index (BMI) of greater than or equal to 30 (12). Symptom and demographic data were complete; comorbidity data were complete insofar as patients had prior encounters in the integrated health system. Mortality data was captured via an existing linkage to state death records.

We used a random number generator to divide the cohort into a 70% derivation cohort and 30% validation cohort. In the derivation cohort data, we fitted a multivariable logistic regression model for hospitalization within 14 days of testing, using clinical and demographic features. Predictors were prespecified before model development based on: clinical features that would be available at the time of testing for all ambulatory and emergency department patients regardless of testing venue (a criterion that precludes, for instance, laboratory data), biological plausibility of association with severity, and reproducibility in other studies in existing COVID-19 literature. We intentionally did not fit a model for mortality, but instead planned *a priori* to validate the ultimate model against that outcome. Model discrimination was evaluated using the area under the receiver-operator characteristic curve (AUROC) and model fit by evaluating R^2^ using the Nagelkerke method (13,14).

We included patients who tested in the ambulatory setting as well as patients who tested positive in the emergency department to ensure that the score would be applicable in both environments. However, we recognized that some patients testing positive in the emergency department (ED) are then subsequently admitted. The decision to admit or not is not immediately known to emergency medicine providers who may still wish to use the score to stratify risk to aid in clinical decision making and selection of therapies. However, because patients who are admitted from the ED may have different characteristics than those tested in the ambulatory setting, we planned *a priori* to perform a sensitivity analysis by repeating the regression above after restricting the cohort to patients who were not admitted to the hospitalization at the time of their test.

We then adapted the original logistic regression model into a simple scoring tool by converting exponentiated β coefficients into weighted point assignments for each variable. We evaluated the test performance characteristics of this simplified clinical prediction tool in the derivation and validation cohorts using AUROC and by calculating the sensitivity, specificity, negative and positive predictive values across the range of scoring thresholds.

## Results

From March 25 through October 1, 2020, 22,816 patients had a positive PCR test for SARS-CoV-2. The mean age was 40 years (see Table 1); 11,424 (50.1%) patients were female and 8753 (43.9%) identified as a member of a community of color (either non-white race or Hispanic or Latinx ethnicity). Patients had on average one significant medical comorbidity. 1419 (6.2%) of patients were admitted; of these, 799 (3.6%) tested positive in the emergency department during the encounter that culminated in admission. Overall 93 patients (0.4%) died within 28 days of their positive SARS-CoV-2 assay. Demographic and clinical features were very similar between derivation (n=16,030) and validation (n=6786) cohorts.

**Table 1.**
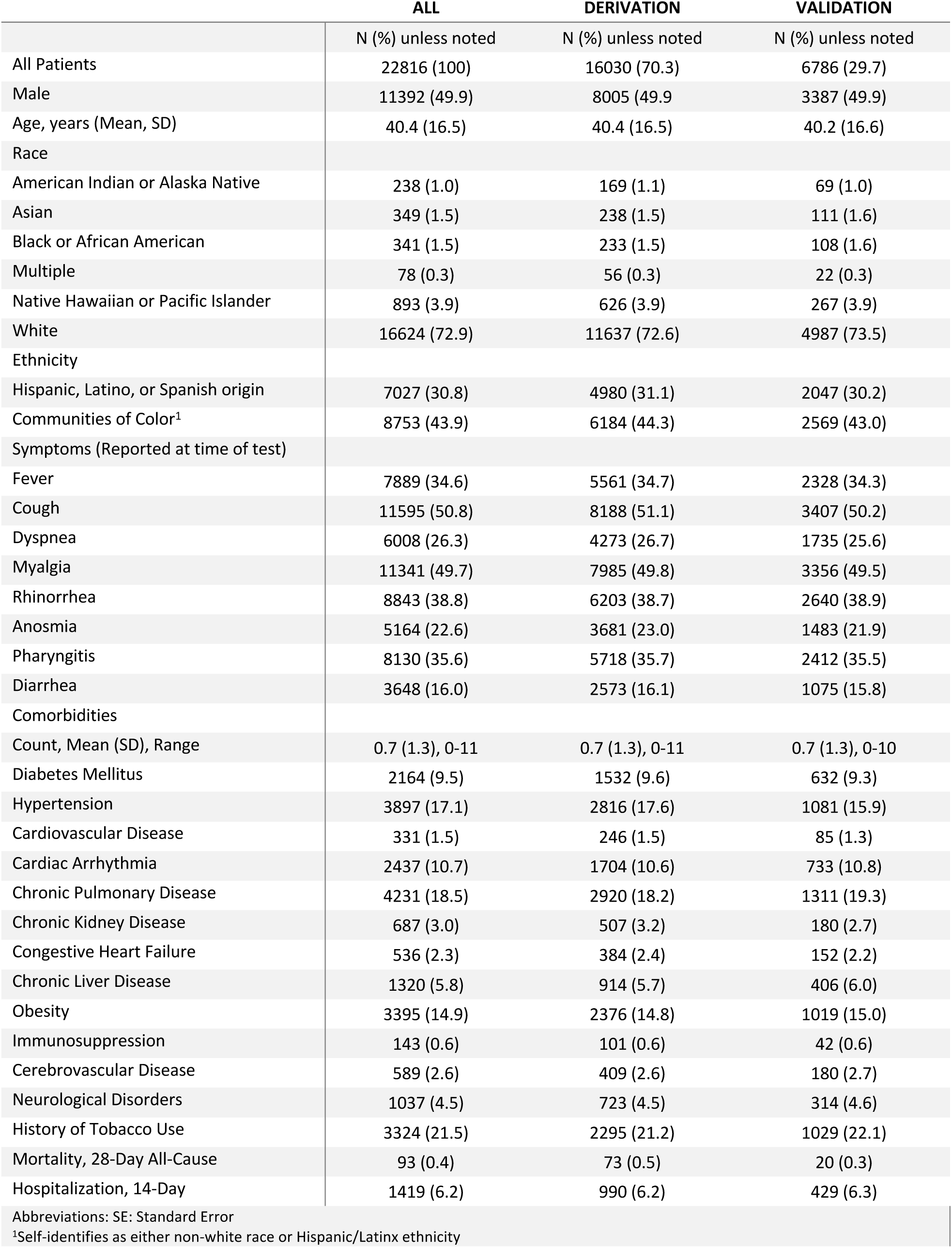
Patient characteristics, total and by derivation and validation cohorts.

**Table 2.** Patient characteristics of the derivation cohort stratified by outcome of hospitalization.

|  | Hospitalized |  |
| --- | --- | --- |
|  | No | Yes |
|  | N (%) unless noted | N (%) unless noted |
| **N=** | 15040 | 990 |
| **Male** | 7472 (49.7%) | 533 (53.8) |
| **Age, Years (Mean, SD)** | 39.5 (16) | 54.8 (17.7) |
| **Race** |  |  |
| American Indian or Alaska Native | 145 (1.0) | 24 (2.4) |
| Asian | 218 (1.4) | 20 (2.0) |
| Black or African American | 217 (1.4) | 16 (1.6) |
| Multiple | 55 (0.4) | 1 (0.1) |
| Native Hawaiian or Pacific Islander | 524 (3.5) | 102 (10.3) |
| White | 10940 (72.7) | 697 (70.4) |
| **Ethnicity** |  |  |
| Hispanic or Latinx | 4622 (30.7) | 358 (36.2) |
| Communities of Color<sup>1</sup> | 5671 (43.5) | 513 (54.7) |
| **Symptoms (at time of testing)** |  |  |
| Fever | 4999 (33.2) | 562 (56.8) |
| Cough | 7575 (50.4) | 613 (61.9) |
| Dyspnea | 3707 (24.6) | 566 (57.2) |
| Myalgia | 7462 (49.6) | 523 (52.8) |
| Rhinorrhea | 5961 (39.6) | 242 (24.4) |
| Anosmia | 3516 (23.4) | 165 (16.7) |
| Pharyngitis | 5476 (36.4) | 242 (24.4) |
| Diarrhea | 2379 (15.8) | 194 (19.6) |
| **Comorbidities** |  |  |
| Comorbidity Count, (Mean, SD), Range | 0.7 (1.2), 0-10 | 2.1 (2.0) 0-11 |
| Diabetes Mellitus | 1145 (7.6) | 387 (39.1) |
| Hypertension | 2308 (15.3) | 508 (51.3) |
| Cardiovascular Disease | 178 (1.2) | 68 (6.9) |
| Cardiac Arrhythmia | 1442 (9.6) | 262 (26.5) |
| Chronic Pulmonary Disease | 2620 (17.4) | 300 (30.3) |
| Chronic Kidney Disease | 357 (2.4) | 150 (15.2) |
| Congestive Heart Failure | 261 (1.7) | 123 (12.4) |
| Chronic Liver Disease | 768 (5.1) | 146 (14.7) |
| Obesity | 1987 (13.2) | 389 (39.3) |
| Immunosuppression | 82 (0.5) | 19 (1.9) |
| Cerebrovascular Disease | 315 (2.1) | 94 (9.5) |
| Chronic Neurological Disease | 573 (3.8) | 150 (15.2) |
| **History of Tobacco Use** | 2012 (20.5) | 283 (28.6) |
| **Mortality, 28-Day All-Cause** | 13 (0.1) | 60 (6.1) |

Abbreviations: SE: Standard Error
<sup>1</sup>Self-identifies as either non-white race or Hispanic/Latinx ethnicity

In the derivation cohort, the primary multivariable model (see Table 3) demonstrated adequate model diagnostics [AUROC 0.824 (95% CI 0.809-0.840), Nagelkerke R^2^ 0.26]. Age, male sex, self-identification to a community of color, dyspnea and high-risk comorbidities including diabetes mellitus, obesity, immunosuppression and chronic neurologic disease were each associated with significantly greater odds of hospitalization. In an exploratory analysis in which individual comorbidities were replaced in the regression with a count of total comorbidities, the cumulative number of comorbidities was also significant (OR 1.4, 95% CI 1.3-1.5). In the planned sensitivity analysis excluding patients who were tested in the emergency department during their admission to the hospital, the multivariable model had slightly diminished performance [AUROC 0.789 (95% CI: 0.768-0.810), R^2^ 0.164]. Overall, contributions of individual risk factors were similar in this model compared to the model including patients being admitted, (see Table 4) with the exception that the magnitude of risk of dyspnea was less in the ambulatory-only cohort (OR 2.1 vs 3.5), and the odds of immunosuppressed patients without palliative goals of care being admitted were greater (OR 7.0 vs 3.9).

**Table 3.** Multivariable logistic regression model for hospitalization in the derivation cohort.

|  | p | Adjusted Odds Ratio | 95% CI |
| --- | --- | --- | --- |
| Age (decades) | <0.0001 | 1.5 | 1.4-1.6 |
| Male | <0.0001 | 1.3 | 1.2-1.6 |
| Communities of color<sup>1</sup> | <0.0001 | 2.1 | 1.8-2.4 |
| Dyspnea | <0.0001 | 3.5 | 3.0-4.0 |
| Diabetes mellitus | <0.0001 | 2.2 | 1.8-2.6 |
| Hypertension | 0.001 | 1.4 | 1.1-1.7 |
| Coronary Artery Disease | 0.45 | 0.88 | 0.61-1.3 |
| Cardiac Arrhythmia | 0.41 | 1.1 | 0.9-1.3 |
| Chronic Pulmonary Disease | 0.39 | 0.92 | 0.8-1.1 |
| Chronic Kidney Disease | 0.29 | 1.1 | 0.9-1.5 |
| Congestive Heart Failure | 0.07 | 1.3 | 1.0-1.8 |
| Chronic Liver Disease | 0.98 | 1.0 | 0.8-1.2 |
| Obesity | <0.0001 | 1.9 | 1.6-2.3 |
| Immunosuppression<sup>2</sup> | 0.02 | 3.9 | 1.3-12.1 |
| Cerebrovascular Disease | 0.74 | 1.1 | 0.8-1.4 |
| Chronic Neurologic Disease | <0.0001 | 1.8 | 1.4-2.4 |

<sup>1</sup>Self-identifies as either non-white race or Hispanic/Latinx ethnicity<sup>2</sup>Excludes patients with metastatic cancer with non-hospitalization goals of care**Table 4. Sensitivity Analysis: multivariable logistic regression model for hospitalization in the derivation cohort, excluding patients admitted from the emergency department**

**Table 4.** Sensitivity Analysis: multivariable logistic regression model for hospitalization in the derivation cohort, excluding patients admitted from the emergency department.

|  | p | Adjusted Odds Ratio | 95% CI |
| --- | --- | --- | --- |
| Age (decades) | <0.0001 | 1.5 | 1.4-1.6 |
| Male | 0.003 | 1.3 | 1.1-1.6 |
| Communities of color<sup>1</sup> | <0.0001 | 1.8 | 1.5-2.2 |
| Dyspnea | <0.0001 | 2.1 | 1.7-2.5 |
| Diabetes mellitus | <0.0001 | 2.1 | 1.6-2.6 |
| Hypertension | 0.001 | 1.2 | 1.0-1.6 |
| Coronary Artery Disease | 0.91 | 1.0 | 0.6-1.5 |
| Cardiac Arrhythmia | 0.39 | 1.1 | 0.9-1.4 |
| Chronic Pulmonary Disease | 0.12 | 1.2 | 1.0-1.4 |
| Chronic Kidney Disease | 0.89 | 1.0 | 0.7-1.4 |
| Congestive Heart Failure | 0.72 | 1.1 | 0.7-1.6 |
| Chronic Liver Disease | 0.87 | 1.0 | 0.8-1.4 |
| Obesity | <0.0001 | 1.8 | 1.5-2.3 |
| Immunosuppression<sup>2</sup> | 0.003 | 7.0 | 2.0-24.9 |
| Cerebrovascular Disease | 0.25 | 1.2 | 0.7-1.5 |
| Chronic Neurologic Disease | 0.81 | 1.0 | 1.4-2.4 |

<sup>1</sup>Self-identifies as either non-white race or Hispanic/Latinx ethnicity<sup>2</sup>Excludes patients with metastatic cancer with non-hospitalization goals of care

Criteria included in the probabilistic, simplified clinical prediction score are displayed in Table 5. Because cumulative comorbidity count was significantly associated with poor outcomes, we included comorbidities in the simplified tool that were not statistically significant individually in the expanded logistic regression model. In the derivation cohort, the AUROC for the simplified clinical prediction score for 14-day hospitalization was 0.82 (95% CI: 0.81-0.84) and 0.8 (95% CI: 0.78-0.82) in the validation cohort. AUROC for 28-day all-cause mortality in the derivation cohort was 0.91 (95% CI: 0.83-0.94) and in the validation cohort 0.80 (95% CI: 0.69-0.9). The scoring threshold that optimized sensitivity and specificity (by Youden’s index) was 6 with test characteristics of 71.1% and 76.2% respectively (Table 6)(15).

**Table 5.** Simplified Clinical Prediction Score for COVID-19 Outcomes.

| Demographic Risk Factors | Points |
| --- | --- |
| Male | 1 |
| Age | 0.5 for every decade:<br>0-10=0.5, 11-20=1, 21-30=1.5, 31-40=2, 41-50=2.5, 51-60=3, 61-70=3.5, 71-80=4, 81-90=4.5, 91-100=5, >100=5.5 |
| Communities of color<sup>1</sup> | 2 |
| **High Risk Comorbidities** |  |
| Diabetes Mellitus | 2 |
| Severely Immunocompromised<sup>2</sup> | 2 |
| Obesity (BMI>30) | 2 |
| **Other Comorbidities** |  |
| Hypertension | 1 |
| Coronary Artery Disease | 1 |
| Cardiac Arrhythmia | 1 |
| Congestive Heart Failure | 1 |
| Chronic Kidney Disease | 1 |
| Chronic Pulmonary Disease | 1 |
| Chronic Liver Disease | 1 |
| Cerebrovascular Disease | 1 |
| Chronic Neurologic Disease | 1 |
| **Symptom Risk Factor** |  |
| Dyspnea | 1 |

<sup>1</sup>Self-identifies as either non-white race or Hispanic/Latinx ethnicity
<sup>2</sup>Solid Organ or Bone Marrow Transplant, AIDS, Active Chemotherapy, or Inherited Immunodeficiency

**Table 6.** Risk Score test characteristics across thresholds.

| Point Threshold | Sensitivity | Specificity | PPV | NPV | % of Positives |
| --- | --- | --- | --- | --- | --- |
| 3 | 95.0% | 28.5% | 7.5% | 98.9% | 72.8% |
| 4 | 89.1% | 45.7% | 9.3% | 98.5% | 56.3% |
| 5 | 80.6% | 62.8% | 12.1% | 98.1% | 39.8% |
| 6 | 71.1% | 76.2% | 16.6% | 97.5% | 26.7% |
| 7 | 60.9% | 84.1% | 20.6% | 97.0% | 18.7% |
| 8 | 51.4% | 89.2% | 24.4% | 96.4% | 13.4% |
| 9 | 41.4% | 92.8% | 28.2% | 95.9% | 9.4% |
| 10 | 32.3% | 95.2% | 31.7% | 95.4% | 6.5% |
| 11 | 25.0% | 97.0% | 36.1% | 94.9% | 4.4% |
| 12 | 17.4% | 98.1% | 38.5% | 94.6% | 2.9% |

## Discussion

Given recent straining hospital volumes and the emergence of promising but limited-availability outpatient therapies for COVID-19, methods are needed to identify patients with COVID-19 at highest risk of progression to severe disease, hospitalization and death. Here we describe a simple scoring model capable of accurately risk stratifying ambulatory and emergency department patients for COVID-19 for subsequent hospitalization and mortality.

One of the primary strengths of this model is the simplified and easily calculable score using features that are widely accessible. In particular, our score does not require laboratory studies, which are unavailable in the majority of ambulatory patients testing positive for SARS-CoV-2. While preserving discriminative value, this simple scoring system has potential to facilitate more widespread clinical application in settings lacking robust integration of informatics. The model was derived and validated in a very large and diverse population in the western United States and is based on risk factors for severe disease that are largely conserved across global populations, including age (12), male sex, overall comorbid burden (13), and shortness of breath at the time of risk stratification. These factors align closely with those included in models derived in other locations and populations (3,5–9,16,17). As a result, we expect that this tool will be generalizable.

Although race and ethnicity are often omitted from clinical prediction models to prevent illegal or unethical profiling behavior, the National Quality Forum recommended that when applications of risk prediction include patient selection for preventive or therapeutic modalities, omission of race or ethnicity can actually cause inequity in healthcare access and worsen outcomes disparity by underestimating risk using other demographic and clinical features alone.(16) In COVID-19, it is now well-recognized that significant outcome differences among communities of color exist with respect to severe illness and hospitalization(19) despite adjustment for age, gender and underlying medical conditions.(5,6) This remains poorly understood and may be due to social determinants of health, inadequate access to healthcare, or poorly-controlled co-morbidities. Because we anticipated application of this risk stratification model to aid in allocating preventive therapies in COVID, we, like other published models(5,6), chose to include race and ethnicity in our score. In future work, more refined socioeconomic, cultural and healthcare access surrogates would be preferable alternatives.

When emergency use authorization (EUA) was granted by the United States Food and Drug Administration for monoclonal antibodies bamlanivimab and casirivimab/imdevimab for administration in non-hospitalized patients with early mild-moderate COVID-19, most states were experiencing peak community transmission, with thousands of new patients per day. It became clear that not only would the supply of drugs be inadequate initially to treat all patients qualifying under EUA criteria, but the capacity to administer infusions without compromising infection control in infusion sites would be even more limited. To address this limited resource situation, the Utah Crisis Standards of Care scarce medications committee was convened with the goal of equitably and efficiently matching available infusion capacity to patients at highest probability of hospitalization most likely to benefit. The simple scoring tool described herein was ultimately adopted because of the simplicity, widely accessible clinical features and validation in a large, representative local population (20). By regularly adjusting the eligibility criteria based on the risk score threshold that best calibrates current infusion capacity to the number of new cases in high-risk strata, this risk-targeted drug allocation strategy has provided an equitable and flexible means of drug delivery in the context of still-uncertain efficacy and limited resources.

Limitations of our study include the retrospective, observational design, and the possibility that comorbidity data may have been unavailable or out of date for some patients in the cohort who receive the majority of their medical care outside our integrated healthcare system. Although the large study population and inclusion of widely recognize features improves the likelihood of generalizability, this will need to be confirmed through external validation.

In this large retrospective cohort study, we identified simple risk factors that can easily be calculated at the bedside without laboratory values to risk stratify COVID-positive individuals for risk of hospitalization and death. Applications include guiding allocation of therapies that are limited in availability. External validation is needed to confirm generalizability in diverse and geographically independent population.

## Data Availability

In order to protect patient privacy and comply with institutional data use policy, data used in this study are unavailable to upload to public servers. As required by the Intermountain Healthcare Institutional Review Board, data sharing agreement requests to access deidentified versions of the datasets generated and/or analyzed during the current study may be addressed to the Intermountain Office of Research

## Author Contributions

Study concept: BJW, JB, IP, PJ, DH, BH, AS, NS, WB, EH, DM, RS, SMB

Study design: BJW, JB IP, SMB, GS

Data collection: BJW, NG

Statistical analysis: BJW, JB, IP, SMB

Interpretation of results: All authors

Manuscript preparation: All authors

Critical review of the manuscript: All authors

